# Eat, Pray, Work: A meta-analysis of COVID-19 Transmission Risk in Common Activities of Work and Leisure

**DOI:** 10.1101/2020.05.22.20110726

**Authors:** Meher K. Prakash

## Abstract

**BACKGROUND:** When the lockdowns are relaxed, the responsibility of mitigating the COVID-19 spread shifts from the governments to the individuals. To know how to conduct one-self, it is important for everyone to know the risks of transmission during the quotidian activities - meetings, meals, etc, from individuals who are known to them and looking healthy.

**METHODS:** The detailed case-studies corresponding to 425 infections upon point-exposures over a specified duration are curated. The data from the case studies is summarized and reorganized to reflect different situations from the daily life. A meta-analysis of the attack rates of transmission and the number of infections per infected person are performed.

**RESULTS:** The attack rates are very high in family dinners (66.7% (48.8-80.8%)) compared to sit-down dinners with lesser mixing among people eating at different tables (15.7% (12.1-20.1%)), both lasting a couple of hours. In an open workspace office floor organized in a two-half structure with shared elevators and restrooms and the employees speaking continuously, the average attack rate over the course of a few days was much higher in one half (78.7% (70.3-85.3%)) than the one for the entire floor (43.5% (37.0-50.1%)). Inferred data suggests that the transmission in elevators and trains may be lower under the conditions of using masks. In most of the instances we studied, the infected individuals spreading (35/44) and even super-spreading (3/6) were mostly without symptoms of coughing, sneezing or a fever.

**CONCLUSIONS:** Although the basic reproduction number R_0_ is around 3.0, the number of infections caused, including the super-spreading events, seem to be limited by the number of personal interactions in a group and their proximity. By acknowledging the risks in daily life, from healthy-looking persons, one may be able to organize their interactions better to reduce the chances of spreading or super-spreading infections.

## INTRODUCTION

For the past few months, many individuals felt at war with the government, as if the policies for COVID-19 mitigation did not concern them. With an intent to restart the economies, the mitigation measures are being relaxed, despite the fears of a second wave. As the individual desire to go back to “normal life” grows stronger, the responsibility of mitigating shifts from the government to the individuals. Although the epidemiologists underscore that one should embrace the “new-normal”, most of the community neither understands nor appreciates what it is. In the past months since the breakout of the COVID-19 pandemic, everyone has been hearing two things very regularly, a very high number of global infections (which is more than 5 million at this stage) and that one should practice social distancing to mitigate these effects [1]. At times one also hears positive reports about containment from the epidemiologists that the R_0_, the basic reproduction number that signifies how many new infections can be caused by an infected person, is reducing due to social distancing measures.

Among all these average descriptors and predictive models, one does not have a feeling for how the infections spread and how high the chance of COVID-19 transmission is. Since there is no vaccine in sight, no slowing down of COVID-19 infections in hot countries, one has to acknowledge the nature of the disease and its transmission and learn to live the new-normal. The transmission risks in person-to-person interactions have been graded by the Centers for Disease Control and have served as general guidelines [2]: living in a same household as an intimate partner of a COVID-19 positive patient (high risk), being within 6 feet (or around 2 meters) from a symptomatic laboratory-confirmed person (medium risk), being in the same indoor environments including classrooms, hospital waiting rooms for a prolonged period with no close contact (low risk).

As one tries to translate these recommendations and apply them to their own daily lives, clearly, visiting wet markets with live animals or patients hospitalized with respiratory illnesses indicate the highest level of risk. But barring those exceptional situations, how much risk of COVID-19 transmission lies in common activities one performs, a question that sounds pessimistic in the first pass. If an acquaintance is not sneezing or coughing is there a risk in meeting with them in close proximity? Even if one person is infected at a party, if R_0_ of COVID-19 is around 3 [3], is there a chance of more than 3 new infections? Some of the studies attempted to perform an overall assessment of primary and secondary attack rates [4] or classify the total number of infections by profession [5], which is very different from the goal we wish to accomplish about the chance of transmitting infections during different daily activities.

Answering these questions requires in a way to redesign daily interactions requires a detailed investigation of the circumstances and activities under which COVID-19 spreads. The information pertaining to the spread and containment of COVID-19 infections is usually available at three levels of granularity:

### Societal level

The total number of infections in a state, or a country has been a commonly reported metric. As countries risked overwhelming of health care resources, there have been several recommendations for the containment of the infections using lockdowns [6]. Epidemiological data [7] where the exponential growth reduced in intensity to a sub-exponential growth confirmed the efficacy of lockdowns. More interestingly, a detailed town-wide testing of the entire village of Vo (Italy) [8] on multiple occasions following a lockdown clearly demonstrated that if the individuals are isolated, new infections outside households can be stopped.

### Transmission clusters

Honing in one level finer, one hears about an incident or a person that triggered a series of infections, dubbed as a cluster. For example, a cluster of around 5000 infections arose in multiple transmission generations, from an infected person attending a Church in South Korea. As the first wave of the pandemic in South Korea comes to an end, this cluster still accounts for more than 45% of the overall infections. A few contact tracing studies provide a closer look into the contacts, such whether it happened during a meal, at a church or in a household [4] or have detailed the number of infections at different workplaces [5]. However, much of the information publicly known information is available at a ‘topological level’ in the language of graph theory, meaning person A infected person B after a “contact”. The contact could mean a continuous exposure, such as two people living together in the same household, known as the familial clusters, or a point-exposure which signifies a specific interaction instance over a well-defined of time spent together.

### Individual level

At the extremely detailed level, many scientific studies focused on the individual case records of the patient-0 or the index-patient – to understand instances of local transmission or to summarize presenting their clinical and virological characteristics [9] as well as to compare the mutations.

Clearly, individual case-studies are the very personal and educational. Our goal in this work is to gain insights from the handful of documented transmission events, among the 5 million infections, to assist in assuming a sense of individual responsibility or help modelling efforts. However, compiling and analysing the infection transmission records is not easy, as many do not know where they contracted the infections from, because there are too many infections in town or because their transmitter was asymptomatic [10] and others who know how they contracted the infection may consider it a violation of their privacy to disclose it.

In early days of COVID-19 infections, many countries investigated and even published the detailed of records that attempted to understand whether the cases were imported from abroad, or were a result of a local transmission. In countries such as South-Korea or Singapore which have performed extensive contact tracing, and detailed transmission chains are constructed. However, the publicly available transmission accounts still date back to the early days of infections, along with some recent accounts of super-spreading individuals or events. We consolidate the information that is publicly available to weigh the risks of transmission in daily life.

## DATA CURATION

### Inclusion Criterion

A literature search was performed in Google, medRxiv and Web of Science, using the key words “index patient”, “Case patient”, “Patient-0”, “COVID-19”, “SARS-Cov-2”. The search was further refined with a special emphasis on articles published in CDC, Lancet, NEJM. In the instances (Itaewon cluster, birthday parties in Pasadena and Connecticut) where the articles were in news media rather than the scientific articles, the data was substantiated using press releases from the governments and alternative sources. The data was included into our analysis if a detailed case history with a point source of infection and a well-defined duration of exposure could be identified. Detailed case studies from the countries where contact tracing was employed: Singapore and South Korea were used as well.

### What is not in scope

If within the selected articles, if the details of the quality of the interaction or the duration of the stay together were not described or cannot be inferred they were excluded. This applied to exclusion of articles as well as exclusion of some cases within the selected articles from which household transmission cases were excluded.

Indirect transmissions, either through unknown contacts and asymptomatic individuals or possibly through fomites [11] and aersols [12], where a clear interaction between individuals could not be identified were also excluded.

In a sense a hospital is a high risk environment. At least two index cases in our curated data contracted the infection while visiting their respective relatives in hospitals [13,14]. At the same time, no secondary infections were caused by a patient in 53 bed ward, to any of the 71 staff and 49 patient contacts (seven staff and 10 patients considered ‘close contacts’) [15]. The possible reasons in this case are that the patient had been symptomatic for about a week before the hospitalization and hence has a lower infectiousness [16,17] or the patient remained in oxygenation for 18.5 out of 35 hour stay in the ward, and other contacts had used varying levels of personal protective equipment [15]. Data on the infections in hospitals is thus scarce and hard to interpret unless it is extremely detailed and is not analysed in this work which focuses on everyday activities.

Several cases, which in a sense seem obvious cases of high transmission risk, but carry few recorded details were excluded - travel to Wuhan of a family of 6, of which 2 spent overnight at a Wuhan hospital treating one of their relatives for febrile pneumonia [13]; a person contracting the infection while visiting a hospitalized family member in Wuhan, and eventually transmitting it to her husband with whom she shared meals and bed for many days and the 119 (34%) medium to high risk contacts that the couple had but they neither resulted positive nor are described in detail [14].

There have been many journalistic accounts of the spread of infections in Italy at a Soccer match, but the number of infections to start with, the chances of other sources of infections, and the time when a majority started showing symptoms could not be identified and hence excluded from the study.

Cruise ships and war ships on the other hand present a well-defined duration of isolation from the land, although very long, and present an interesting case, and are treated separately.

## RESULTS

### Case history summaries

The curated data from different articles [18-31] is summarized (**Supplementary Information**) as 20 situations. Where possible, the original labelling used for defining the patients (Patient 0, Patient 1, .. or Patient A1.1, A2.1, ..) has been preserved in order to making the retracing of the accounts in the original articles easy. The case summary situations were further subdivided for classification into different types of instances. Of these 20 situations, 418 infections occurred on 32 instances and caused by 44 infected individuals were identified. These instances or the infections do not include the data from the cruise/ships which was analysed separately. The separate instances from these case-summaries reflecting the different activities one is likely to perform were gathered and analysed in the **Supplementary Tables 1 to 8**. A final summary of the attack rates inferred from these **Supplementary Tables** is presented in **Table 1**. Among all the 44 individuals spreading the infections in these 32 instances, only 8 individuals had mild symptoms and contributed to about 113 of the 418 infections.

**Table 1.**
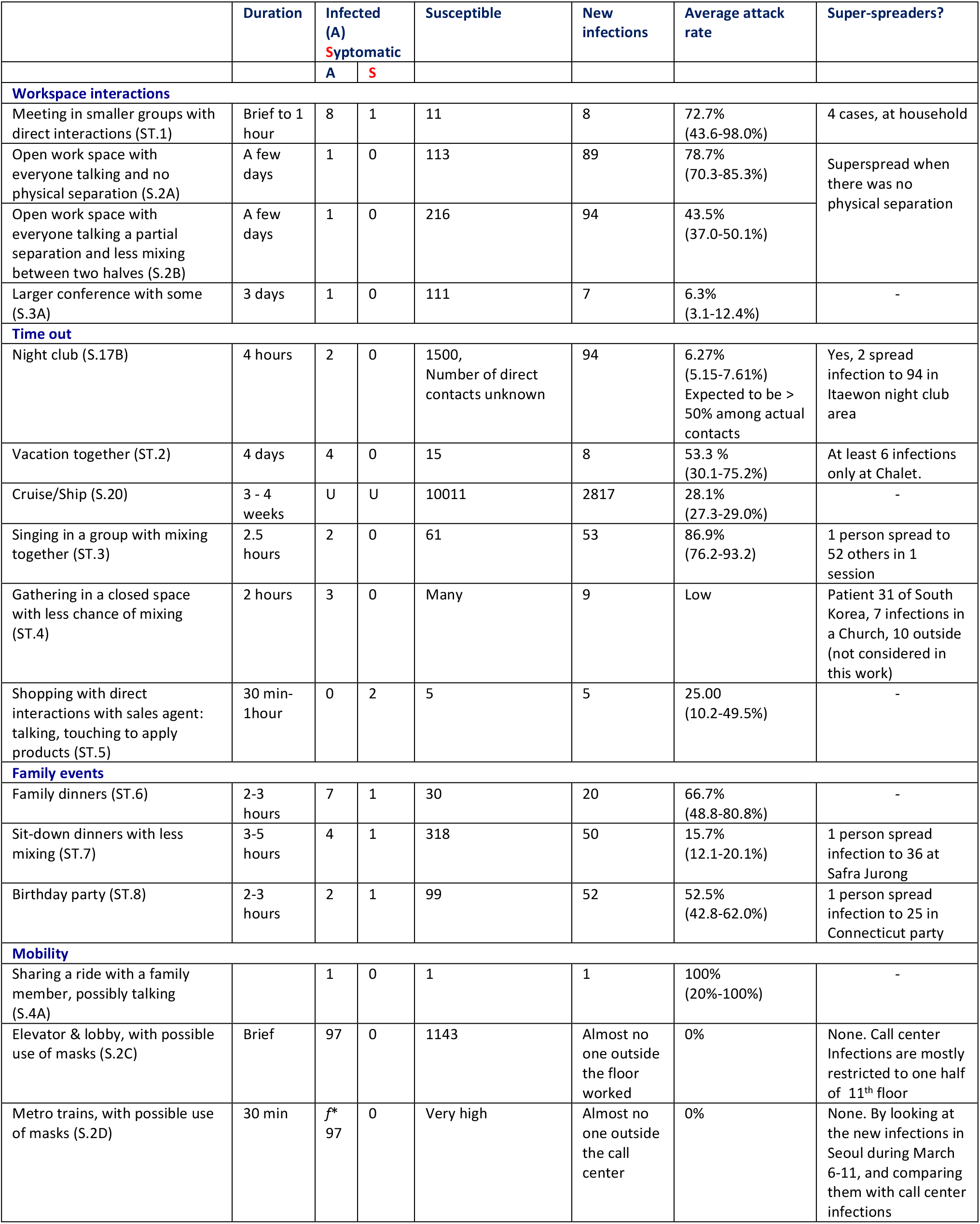
A summary of the transmission risks curated from several sources. Detailed numbers of each category are shown in Supplementary Information. U is unknown. *f* is an unknown fraction, for which there are reasons to believe can be around 0.5 at least. In each case the source of the numbers compiled in the **Supplementary Information** is cited. For example, **ST.1** and **S.20** refer to the supplementary table 1 and Situation 20.

### Transmissions at Meals, Singing and in Meetings

The case studies where we could identify a point-exposure to an infected person, on a specific occasion and for a well-defined duration, center around meals, meetings, and singing events either for recreation or religion. The attack rates are defined as the fraction of susceptible individuals, who after coming in contact with the infected individuals also got infected. The susceptible individuals are the ones that are truly likely to come in contact, such as people working in the same floor or eating at the same table rather than individuals in other floors of a building which is poorly connected. The detailed transmission risks for each scenario are given are categorized under 4 broad heads – work place interactions, activities of leisure, family events, and related to mobility (**Table 1**). Each of these categories has several events with high-risk of transmission. Short private meetings lasting 1 hour in a work place have a very high transmission rate (72.7% (43.6-98.0), comparable to working together on the same open floor where there is sufficient movement of people (78.7 (CI. 70.3-85.3%)). Practicing singing in a group for around 2 hours, along with a high mixing interactions in the group had a very high transmission rate (86.9% (CI.76.2-93.2%)), whereas singing and gathering in very large groups with close interactions with neighbors and otherwise not much mixing (such as in prayer services) resulted in 1-7 infections by several index-patients. Traveling in a car with a family member by talking has a very high risk (100% (CI.20-100%)), while traveling in metro trains with masks and possibly no talking has the lowest risk (~0%).

In quick summary, whenever a high quality of person to person interaction is possible, the chance of spreading infection is much higher than 60%. None of these results go against the standard recommendations of social distancing, however, they clarify the nature and duration of the interactions and quantify the transmission rates.

### Nature of Super-spreaders

In pandemics it is always important to understand how the super-spreaders can be identified and stopped [32]. Among the data analysed, if one gathers the number of infections caused by different people in a given instance, the median of these numbers results to be 2.5, (**Supplementary Table 9**) as expected from the basic reproduction rate (R_0_) of COVID-19 [3]. With this validation, we examine the curated data for the super-spreaders. Super-spreaders is a qualitative term, and in this work we consider anyone spreading more than 3 (~R_0_) infections as a super spreader. Among the 44 individuals who caused infections, six super-spreaders could be identified, which curiously fall in two groups - A. Infections on multiple occasions - Patient 31 in South Korea who infected 7 at the Church (**S.12A**), and 10 others in different occasions and different days (**S.12B**), Patient A1.1 who attended the funeral in Chicago, who caused 3 infections at the funeral (**S.18A**), and 7 others at the birthday party 3 days later (**S.18B**), B. Infections on a single occasion – 6 in the Alpine vacation (**S.10**), 51 Infections caused at the Choir in Washington (**S.14**), 36 infections caused at the dinner in Safra Jurong (**S.6**), and 94 infections caused by the two attended 5 night clubs (**S.17B**). The index patients at the funeral in Chicago, the dinner at Safra Jurong and Choir in Washington had mild respiratory symptoms, and in the other three cases did not have any symptoms when they infected others. In general, various estimates suggest that the infectiousness drops significantly once the symptoms develop [16,17]. However, there may be exceptions and an individual may continue to spread infection on multiple days. At the same time, any person with a common level of infectiousness and without symptoms can become a super-spreader by having very close interactions with many people or actively mixing in large congregations. There was also a situation of a teacher who attended one of the Itaewon nightclubs (**S.17C**) in discussion and spread infection to at least 15 individuals, including 6 students. However, the detailed interactions are not known and hence not analyzed.

By taking a look at the super-spreading events in the curated data, it is clear that number of infections caused in an instance is limited by the number of people one directly interacts with in close proximity, underscoring the possibility that anyone can become a super-spreader if sufficient care is not exercised. This rationale is also the microscopic equivalent which moots the arguments that use per-capita infections across the countries to compare not the overall well-being but rather the rates of infections. Whether it is the spread of infections among the individuals in a gathering or in a country what matter is the number of susceptible people that are truly likely to come in contact with the infected individuals and the quality of those interactions.

### Role of physical separation

Within the data we analysed, one clearly sees the mixing of the people, coming closer together has a strong influence on the spread of infections, much in line with the general social distancing recommendations. Consider the difference in the attack rate between the family dinners (66.7% (CI48.8-80.8)) and birthday parties which can be imagined to be more personal, with stronger physical interactions compared to the sit-down dinners where the people move around much lesser (15.7% (CI12.1-20.1)) (**Table 1**). Similarly, the seating plan from the call center [20], shows a significantly lower number of infections (5) on one side relative to the other side (79) (**S.2A**), and of course, mostly the infections (94/97) are restricted to the 11^th^ floor which is inhabited by 216 out a 19 floor building with 1143 occupants (**S.2E**). The physical separation draws an interesting interpretation in the case of the spread of infections in a Chinese restaurant [22], where the attack rate among all attendees to the restaurant is much lower (9.9% (CI: 5.3-17.7%) (**S.7A**) than that for the individuals seated along the line of the air-flow as per the graphic in the draft 45.0% (25.8-65.8%) (**S.7B**)(however, considering that no negatives from families A,B,C were reported there is a chance for interpreting it as even higher).

### Transmission in transports and possible role of talking

The only known instance of a family member driving together, and passing on the infection (**S.4A**) clearly shows that proximity in a closed environment and possible talking play an important role. There could be many reasons behind it such as loud talking or singing which can emit aerosols [33] comparable in amount to coughing as was seen in the spread of tuberculosis [34] and assist the spread of COVID-19 [35]. But such analysis is out of the scope of this work. The present work focused on the known case histories of people interacting with others, either at home or at work, and one common theme among all these interactions is talking, even more than any documented evidences of physical contact through hand-shakes. Thus, by construction the data chosen does not have situations where people did not talk. However, two pieces of data are worth inferring from the study on the South Korean call center. Although the elevators were shared by individuals from all floors, the infections that spread over a few days were confined mainly to floor number 11 (**S.2A, S.2E**). Secondly, the call center in South Korea is close to two major metro stations, and many employees use these services. There was a chance that the infections could have spread to people sharing the metro services, which is not an easy to track data. However, the first infection in the call center was reported on March 8, and the total number of infections from Seoul in the March 7-March 14 is 139, which leaves at most 42 infections outside the 97 infections related to the call center cluster. Given the numbers of thousands of susceptible people that will be exposed in the metro trains to a significant fraction (considered to be roughly 0.5×97 for the sake of this discussion), 42 infections even if they were all assigned to the call center cluster still suggests a very low attack rate. Thus, the attack rate in the elevators and the metro trains may be considered almost negligible under the conditions the South Koreans usually travel with masks.

### Symptomatic versus asymptomatic infectees

An infrared scan of temperature was commonly used to quickly screen for COVID-19 patients. However, in many occasions (Safra Jurong dinner in Singapore (**S.6**), night club cluster in South Korea (**S.17**)) there is documented evidence that every visitor was asked the mandatory questions about their travel history and their temperature was checked and yet infections spread. In the data we analysed, only on 6 instances index patients showed mild symptoms compared to 38 instances where the index patient did not show any symptoms of cough, sneezing or fever until a few days later. As noted earlier, even among the super-spreaders 2 were symptomatic, and 4 asymptomatic, which is an important warning against underestimating the risks from seemingly healthy individuals. This argument which aligns with the estimation that 44-68% of infections are passed on in the pre-symptomatic phase [16,17].

### Cruises

Time spent on a cruise or a ship partly qualifies for our eligibility criteria – the vacation is long and lasts a few weeks, and not a common activity that fits naturally into the daily activities. However, during this long stay on board, there are no new sources of infection as it is completely isolated from the coast, and the infections continue to spread starting with a mostly unknown number of index patients. Regardless, since the data from three different different ships, 2 of defense and 1 of vacation was available (**S.20**), we analysed it. Over all, the attack rate in the few weeks is 28.1% (27.3-29.0%), which is surprisingly much lower than the infection rate in most of the personal interactions we studied in this work. The passengers/staff may have been segregated in different decks which may have slowed the infection rate.

### Unique instances

Among the data analysed, there are several other daily situations which are very unique. One would like to have more studies such as: infection spread in a working space which is not a call center and hence did not involve too much talking, transmission or lack of it when driving together in a car with and without talking; the spread of infections in a class room setting, etc. Although there are reported instances of teacher in Itaewon cluster who spread infections to 15 including of his students (**S.17C**) and dancing event at the dinner where the lion dance troupe clothed in costumes with no new infections (**S3.D**), more detailed information is required to evaluate the risks in these situations.

### Conclusions

In conclusion, by curating and categorizing the data according to the daily activities we could quantify the risk of transmission in these different settings. These transmission risks when attending meetings, dinners with healthy-looking infected individuals are surprisingly high. In a sensitive phase when the responsibility of COVID-19 mitigation shifts from the governments to the individuals, the analysis underscores the need not to lower the guard, as a healthy looking normal infected individual can turn out to be a super-spreader if (s)he makes sufficient contacts with large number of individuals in one event.

**Figure 1.**
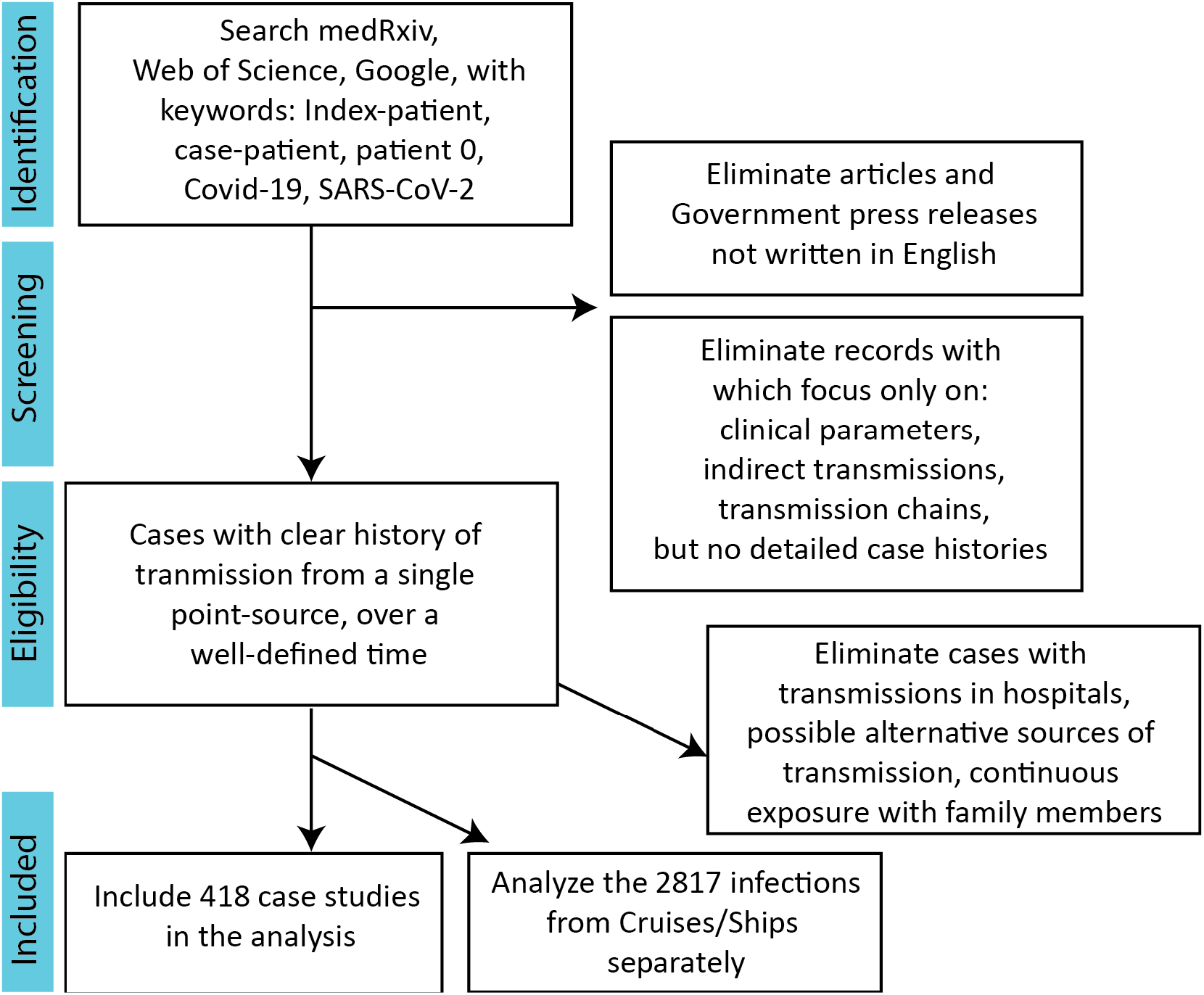
PRISMA Flow diagram for the cases included in the present work.

## Data Availability

Data used has been compiled from public sources. The analysis will be made available upon request

## SUPPLEMENTARY INFORMATION

### Situation 1. Directed Interactions at a workplace. Germany

Total new infections after the event: 16

Ref. Böhmer et al. Lancet 2020 [18]; Rothe et al. N Engl J Med, 2020 [19]

A visitor from Shanghai (Patient 0) attended a training session in Munich during January 20, 21. A few days before this visit on January 16, Patient 0’s parents visited her in Shanghai from Wuhan. During the meetings in Munich she felt fatigue which she thought was due to jet lag, but fever (38.6) started only on the flight back.

S.1A. Patient 0 sat through a 1 hour meeting 3 others in a 12 m^2^ room, including Patient 2 who sat on her side of the table. Only Patient 2 contracted the infection.

S.1B. Patient 2 had a brief contact with Patient 1.

S.1C. Patient 3 and Patient 1 worked simultaneously on the same computer for a short period of time on January 24

S.1D. Patient 12 had a private meeting with Patient 3 for 90 min, and later spent time with Patient 3 and Patient 3’s partner. Partner was negative.

S.1E. Patient 4 sat back to back in a canteen and Patient 5 borrowed salt shaker after asking for it

S.1F. Patient 5 is from a 5-member household and Patients 6,9,11 are from it. Since a specific instance cannot be assigned, these cases are not included.

S.1G. Patient 7 met with Patient 5 for a 1.5 hour meeting at 1.5 meters.

S.1H. Patient 8 and Patient 5 had regular meetings Jan 22, 23, 24.

S.1I. Patient 13 accompanied Patient 0 in travel from China and during the work in Munich.

S.1J. Patient 14 is a household member of Patient 7. This case was not included in our analysis.

S.1K. Patient 7 had a 1 hour meeting with Patients 10, 16.

S.1L. Patient 16 in a meeting with patient 8 > 1 m. Duration not known. Also Patient 16 was also likely to have been infected by 7. Since the case is similar, the infection of Patient 16 is assigned to Patient 7.

### Situation 2. Open-floor work place interactions for a few days, with a lot of talking. South Korea

Total new infections after the event: 97

Ref. Park et al., Emerg Infect Dis, 2020 [20]

In a 19 floor building, has call centers on floors 7,8,9,11 with around 200 employees each. COVID-19 broke out on floors 10, 11. Of the 1,143 (99.8%) individuals including employees, residents and visitors 97 (8.5%, CI 7.0–10.3) tested positive. The first case-patient who worked on the 10th floor and reportedly never went to the 11th floor, had symptoms starting from February 22. The case patient from the 11^th^ floor had symptoms from the 25^th^ of February. The call center came under close surveillance on March 8^th^.

S.2A. According to the graphic given in Park et al., 79 infections were in one half of the 11^th^ floor and only 5 were on the other half (the rest of the 94 cases were not shown in the representation). The two halves shared elevators and restrooms.

S.2B. Total infections on the 11^th^ floor were 94 out of 216.

S.2C. The lobby and elevator by shared by all residents, employees and visitors. Yet the infections were mainly restricted to 11^th^ floor.

S.2D. Metro. The call center is close to two popular metro stations, and many employees use metro services. However, it is hard to trace who all got infected in metro trains. On the other hand the total number of infections reported from anywhere in Seoul during March 7-March 14 was 139, which leaves at most 42 infections that may have been caused by the employees at the call center.

S.2E. Considering all the 1143 (residents, employees, visitors), the attack rate was 97/1143.

### Situation 3. Business conference at a hotel for 3 days. Singapore

Total new infections after the event: 7 Ref. Pung et al. Lancet, 2020 [21]

A British company held an annual 3 day international sales conference from January 20^th^ at a Hotel in Singapore. 94 attendees from 19 countries were among the 111 attendees. 17 attendees were from China, at least 1 from Wuhan. Activities included business presentations, workshops, breakout discussions, team building games which involved close physical contact, city tour. Patient B1 (South Korea), B3 (Malaysia) and B4 (Singapore), B7 (British) were at the same tables for 3 hours at the Chinese dinner (20 January) with approximate 8 member round table. B1, B3, B4, B5 were in the same breakout session of 41 participants for 4 hours, along with 10 Chinese team members.

S.3A. In the entire conference with 111 participants, there were 7 new infections at the end (3 Singapore (3/15), 1 British, 2 South Koreans, 1 Malaysian)

S.3B. Two of the infected members were from one dinner table, and three from another. S.3C. 3 hour workshop where B2, B4, B5 participated. No clear interaction data and hence not used in our analysis.

S.3D. Lion dance troupe spent 5-10 minutes with their lion masks, going around everyone. No personal interaction, did not eat food and did not contract infections.

### Situation 4. Driving Together and Family Dinner

Total new infections after the event: 2

Ref. Pung et al. Lancet, 2020 [21]

The Malaysian attendee to the business conference in Singapore (Situation 3 above) had come in contact with including, family, friends, and medical staff. He caused 2 new infections.

S.4A. He rode in a car with his sister (which we infer as including speaking and sitting side by side)

S.4B. He shared Chinese New Year meals with family January 26, 27,28. His mother-in-law contracted infection. The total number of attendees is not known.

### Situation 5. Family dinner for a few hours at Mei Hwan Drive, Singapore

Total new infections after the event: 7

Ref. Press release from the Ministry of Health, Singapore, Dated 25 February.

https://www.moh.gov.sg/news-highlights/details/links-established-between-church-clusters-and-wuhan-travellers

On January 25, there was a small family get together for celebrating Chinese New Year on the Mei Hwan Drive. This was attended by a couple (Case 83, 91) who also attended a service on January 19 at the Life Church.

S5. Nine of the attendees including Patient 66 (who later spread to Grace of God church), 70, 71, 80, 83, 84, 88, 91 got infected at the incident. Whether there was anyone at the dinner who did not contract the disease is not clear.

### Situation 6. Large Sit-down Dinner Gathering. Safra Jurong, Singapore

Total new infections after the event: 36

Ref. Press release from the Ministry of Health, Singapore, Dated 8 March.

https://www.moh.gov.sg/news-highlights/details/eight-more-cases-discharged-eight-more-cases-of-covid-19-infection-confirmed

Around 200 guests attended a sit-down dinner at Safra Jurong on February 15^th^. As per the regulation, the the attendees were checked for fever, and enquired about a travel history to China in the 14 days before that. All attendees appeared safe as per these criteria, although the Government later announced that there was one person who had symptoms, possibly not fever, attended the event. The dinner, which featured song-and-dance performances and lasted for five hours. There were an estimated 30 tables, and the guests moved between the tables for chit-chat and pictures.

S.6 Within a few days, 36 infections of the attendees were identified. However, considering only 20 families were among them, the remaining 16 could have been either primary infections at the event or secondary infections in the household. We infer the data as all primary infections.

### Situation 7. Eating at a Restaurant in Guangzhou, China. Effect of Air conditioning

Total new infections after the event: 9 Ref. Lu et al. Emerg Infect Dis. 2020 [22]

On January 24, a day after returning to Guangzhou from Wuhan, index patient (patient A1) went for a restaurant with 3 other family members (A2-A4). The restaurant is on the third floor of the 5-floor building (145 m^2^) has its own air coniditioner (AC). Two families B and C with no known history of travel to Wuhan were in the restaurant overlapping around 1 hour each with family A. The seating arrangement showed that they were all in the line of air flow and return (AC ⇄ Family C ⇄ Family A ⇄ Family B). Later that day, patient A1 experienced onset of fever and cough and went to the hospital.

S.7A. On January 24, 91 persons (83 customers who ate lunch at 15 tables, 8 staff members) were in the restaurant. Among the 83 customers, 10 (including the index patient) were ill with COVID-19.

S.7B. Among the people that sat for the longest exposure time, along the airflow direction which could have brought about the mixing, by February 5, 9 members had COVID-19 (4 from family A, 3 from family B, and 2 from family C). The minimum distance to family A was around (B) 1 meter in the direction of flow and 2 meter in the direction of return flow (C) respectively.

### Situation 8. Family home dinner for a few hours

Total new infections after the event: 5

Ref. Zhang et al., Critical Care, 2020. [23]

On January 19, Index patient came back to Bejing from Wuhan. He invited his nephew (Patient 3) for a dinner on the same day. Patients 0, 1, 2, live together, and Patients 3, 4 together and all contracted COVID-19.

S.8. In our analysis only the transmission to Patient 3 at dinner was used and other household transmissions were excluded.

### Situation 9. Family home dinner for a few hours in Nanjing, China

Total new infections after the event: 10 Ref. Huang et al. Lancet, 2020. [24]

On Jan 21, 2020, the index patient left Xiaogan, switched a train in Wuhan and arrived in Nanjing, where she stayed with her sisters (patients 1 and 2) and her mother (patient 3). On Jan 21, she had a dinner with her mother, two sisters, and her brother (patient 4). The index patient had another family dinner with eight relatives on Jan 23. At both these dinners the index patient had no symptoms, but developed cough and fever on Jan 28. Three patients (patients 1-3) who lived together with the index patient and three relatives (patients 4, 6, and 7) who attended the dinner with the index patient on Jan 23 were positive for SARS-CoV-2 infection thereafter. On Jan 24, there was another family dinner with 13 relatives, and two patients (patients 6 and 7) who were still without attended it causing three new infections (patients 8–10). Throughout this familial clusters, the index patients were asymptomatic. Household infections 1,2,3,5 are not used in our analysis.

S.9A. Dinner of Jan 23. Patient 1 attended a dinner with 8 family members, resulting in 3 new infections.

S.9B. Dinner of Jan24. Patients 6,7 attended a dinner with 13 relatives resulting in 3 new infections Patients 8, 9, 10.

### Situation 10. Vacation for 3 days in French Alps

Total new infections after the event: 7 Ref. Hodcroft, Swiss Med. Wkly, 2020. [25]

A British citizen flew after the sales training conference in Singapore from 20-23 January (Situation 3) and joined his family and friends for a 4 day vacation in a Chalet in French Alps on January 24. The group skied, dined together and mostly stayed indoors interacting. He flew back to England on the 28^th^ of January.

S.10 Of the 11 people that were identified as the close contacts to the index patient – 5 infections (in total, including a 9-year-old boy) were detected in France, 1 upon returning to Mallorca and the index patient was diagnosed in England in early February.

### Situation 11. Sharing a room for a few days

Total new infections after the event: 1 Ref. Phan *et al*. N. Engl. J. Med, 2020 [26]

A couple returned from a travel to Wuhan (although they were not exposed to the wet markets), and for three nights from January 17, they shared a hotel room with an air-conditioner in Vietnam with their son. The son had no other known exposure to infection.

S.11 By January 20, the son developed dry cough and was later diagnosed as infected.

### Case 12. Attending a gathering for few hours, with too many people and singing

Total new infections after the event: 7

Ref. Kim et al. medRxiv 2020 and the associated data [27]

https://github.com/yejinjkim/covid19-transmission-network/tree/master/data

Press release dated 24 March 2020, By the CDC Korea,

https://www.cdc.go.kr/board/board.es?mid=a30402000000&bid=0030

On February 7, a person who will later be labeled as Patient 31 was admitted to hospital for an accident. On February 9 she attended a 2 hour service at the Shincheonji church, and another similar session on the 16^th^ of February. Altogether 9300 attended these two services. In these services, usually no face masks or glasses were allowed, and usually people sat in very close arrays to reach the shoulders of neighbors, sang for about 30 minutes, and possibly engaged in potluck meals. She developed fever symptoms on the 10^th^ of February, and was announced COVID-19 positive on 18^th^ February. By March 25, more than 5000 cases could be linked to the Shincheonji cluster.

S.12A. Patient 31 was related to 1160 contacts, and was responsible for 7 infections at the Shincheonji church (from the data sheets in the Supplementary Information of Kim et al. 2020)

S.12B. Patient 31 also caused 10 infections outside the Church. (from the data sheets in the Supplementary Information of Kim et al. 2020)

### Situation 13. Closed space gathering for a few hours

S.13A. Life Church cluster in Singapore.

Total new infections after the event: 6 Ref. Pung et al. Lancet, 2020 [21]

On January 19, 142 of the 227 regular church goers attended a 2 hour long service, to which a couple (case 8, case 9) who returned from Wuhan on the same day visited. The result was 6 infections (Case 31, case 33, case 38, case 83, case 90, case 91). No details of where they sat or how they interacted with the two index patients are known.

S.13B. Grace Assembly of God cluster in Singapore

Total new infections after the event: 9

Ref. Ministry of Health, Singapore, Press release

Case 66 in Singapore who attended the Mei Hwan Drive dinner, attended two sessions at the Grace Assembly of God church. The staff devotion meeting on 29 January, where case 51,48,49 were present. At the prayer meeting, on February 5^th^ there were Case 48, 66, 49. And resulted in 3 staff (54,57,60) and 6 visitors (53, 61, 62, 63,67,73) got infected. The total number of attendees is not known.

### Situation 14. Singing together for a few hours. Choir in Washington

Total new infections after the event:52

Ref. Hamner et al., MMWR Morb Mortal Wkly Rep 2020 [28]

On March 10^th^ 61 of the 122 member Choir attended a rehearsal between 6:30 to 9:00 pm. By March 12 at least six members of the group had developed fever and communicated it to its director. The index patient was later identified as someone who had onset of symptoms on March 7. During the practice, the 61 members occupied the seating arrangement of six rows of 20 chairs each at 6-10 inch distance (seating plan not disclosed by the authorities). There are reasons to believe a strong mixing in the group as there was a 40 minutes joint practice session, other sessions of 50 minutes in two smaller groups, snack break for 15 minutes and gathered together to restore the chairs to the racks.

S.14 Among 61 persons who attended a March 10 choir practice at which one person was known to be symptomatic, 53 cases were identified.

### Situation 15. Singing in a closed space

Total new infections after the event: 3

Ref. W. E. Wei, et al., MMWR Morb Mortal Wkly Rep. 2020 [29]

S.15A. On February 15, Patient B1 attended a dinner where she was exposed to a COVID-19 positive patient. On February 24, patient B2 and patient B1 attended the same singing class. Patients B1 and B2 developed symptoms on February 26 and 29, respectively.

S.15B. Patient F1 attended a singing class on February 27^th^, where she was exposed to a COVID-19 positive patient. On March 1 F1 attended a Church service an infected Patients F2, F3 who were one row behind her. Patient F1 and F2, developed symptoms on March 3rd, and F3 on March 5^th^.

### Situation 16. Shopping for a few hours with direct contacts with sales agents. Singapore

Total new infections after the event: 10

Ref. Pung et. al. Lancet, 2020 [21]

A tourist group of 20 visited Singapore on January 22, 23 from Guangxi, China. During the tour five or six were suspected to be coughing, although only two of them later tested positive on January 28 (father and daughter AGX1, AGX2).

S.16A. The tourist guide (AT1) who interacted with them for a day was COVID-19 positive. However, this is a continuous exposure, with a lot of talking for a complete day and hard to track the interactions. Hence we do not consider this case in our analysis.

S.16B. The group visited a jewelry shop for 1 hour, and the agent who interacted with them AJ1 tested positive. AJ1 also happens to be the husband of AT1, however, since the tour spent 1 hour with AT1, we classify this as a potential direct infection.

S.16C. The group also visited a complementary health products shop where the 4 sales agents who interacted with them tested positive. The sales agents in the shop typically apply products on the customers, and usually do not have handwash in between customers. There were 16 staff altogether, but possibly only 4 interacted with the group. S.16D. At the home of the tourist guide AT1, there were two other infections 6-month old child and domestic help. However, the length of interactions not known and hence not included in our analysis.

### Situation 17. Attending night clubs with too many people for a few hours. Itaewon cluster, South Korea

Total new infections after the event: 94 as of May 19

Ref. Information and numbers of infected people from the Press Releases of CDC Korea, May 19^th^.

https://www.cdc.go.kr/board/board.es?mid=a30402000000&bid=0030 Consistent account reconstruction from multiple media reports.

https://www.straitstimes.com/asia/east-asia/s-korea-delays-school-reopening-as-club-cluster-grows

https://time.com/5834991/south-korea-coronavirus-nightclubs/

https://en.yna.co.kr/view/AEN20200509001452315

On April 30, a 29 year old spent a day at the resort with three friends. After returning, he and one of the friends he traveled with, went to party at 5 night clubs between the night of May 1 and early hours of May 2. The club, as per the regulations tested for temperature, asked for travel history and noted their contact details. However, nothing looked suspicious. However, the index patient had a high fever and diarrhoea on the evening of May 3 and tested positive on May 6.

S.17A. Resort stay with 1+3 friends caused 1 new infection. 2 other friends tested negative.

S.17B. The turnout at the clubs was 1500. The press releases mention that 94 infections are caused during May 2-3 among the club-goers, in addition to more than 90 secondary infections. We infer that most, if not, all of these 94 primary cases may be attributable to the couple of friends. The reasons being that they visited they were in 2, and visited 5 different heavily crowded places, which already is likely to create sufficient new infections. The chance that others who were infected on May 2 early hours, became infectious, and visited the clubs again on May 3 early hours makes it less likely that the 94 infections were caused by individuals other than the two friends.

S.17C. One of the persons who was infected at the club is a teacher in Incheon, and subsequently infected at least 15, in which there were 8 middle and high school students, a female middle school student privately tutored by the instructor, her twin brother and their mother. However, the detailed interactions of how this teacher contracted or transmitted infections is not known and hence it is not included in our analyses.

### Situation 18. Funeral followed by a birthday party in Chicago

Total new infections after the event: 15

Ref. Ghinai et al. MMWR Morb Mortal Wkly Rep, 2020 [30]

In February 2020, a funeral was held in Chicago for a person who died of a non-COVID-19 causes. The index patient (A1.1), a close friend of the family, had recently travelled out of the state and experiencing mild respiratory symptoms had attended the funeral.

S18A. The evening before the funeral, patient A1.1 shared a 3 hour long meal, eaten from common serving dishes, with two family members (patients B2.1 and B2.2) of the deceased at their home. At the funeral which lasted about 2 hours and involved a “potluck-style” meal, patient A1.1 to express condolences embraced the family members B2.1, B2.2, B2.3, and B3.1 of the deceased. Patients B2.1, B2.2, B2.3 developed symptoms within 2, 4 and 6 days respectively after the funeral.

S.18B. Three days after the funeral, patient A1.1, attended a birthday party attended by nine other family members at the home of A2.1. Patient A1.1 embraced others and shared food at the 3-hour party, and close contact between A1.1 and all other attendees is reported. Seven party attendees, Patients A2.1 to A2.7 were infected.

S.18C. A few days after the birthday party, patients A2.5, A2.6, and A2.7 attended a Church where they spoke to a healthcare professional ((patient D3.1) with no other known exposure to COVID-19 for 90 minutes, within a 1 row distance and the offering plate was passed. D3.1 was symptomatic within a day.

### Situation 19. Two different birthday parties

Total new infections after the event: 25

Ref. https://www.nytimes.com/2020/03/23/us/coronavirus-westport-connecticut-party-zero.html

S.19A. **Birthday in Connecticut**.

On March 5^th^, 50 guests from across US and abroad attended a birthday party at home in a wealthy suburb in Connecticut. About half of the attendees to this lavish buffet meal were infected. No one had symptoms at the party, the first reported symptoms were by a person who felt sick on the flight back home. The positive cases went back to other cities in USA as well as to other countries.

### Situation 19. Birthday party in California

Total new infections after the event: 20

Ref. https://losangeles.cbslocal.com/2020/05/09/large-birthday-celebration-in-pasadena-traced-to-5-confirmed-covid-19-cases/

An April birthday party in Pasadena with an estimated 30-40 attendees had 5 confirmed cases, many others (> 20 with symptoms). One of the guests at the event was coughing. No masks were used, no social distancing was maintained.

### Situation 20. On board Cruises and Ships, for a few weeks

#### S.20 A. Diamond Princess

Total new infections after the event: 634

Ref. Kenji et al., Euro Surveill. 2020 [31]

An 80-year-old passenger traveled from Yokohama to Hong Kong. Six days after disembarking the ship, on February 1, she tested positive and on February 3, Diamond Princess was put into quarantine. By 21 February, 634 out of 3,711 passengers and crew members on board Diamond Princess cruise tested positive.

#### S.20 B. US aircraft carrier Theodore Roosevelt

Total new infections after the event: 1,102

Ref. https://www.navytimes.com/news/your-navy/2020/05/06/navy-puts-a-halt-to-daily-covid-19-updates-for-stricken-ships-theodore-roosevelt-and-kidd/

Theodore Roosevelt in early March had on board a few sailors that stayed in a hotel in Vietnam where two people tested positive. After staying at sea for more than a month, 1,102 out of the total crew of 4,500 tested positive.

#### S.20 C. The French navy’s Charles de Gaulle

Total new infections after the event: 1,081

Ref. https://www.navytimes.com/news/your-navy/2020/04/20/french-carrier-surpasses-theodore-roosevelt-with-over-1000-confirmed-cases-of-covid-19/

Charles de Gaulle was at sea for more than a month, and 1,081 of the nearly 1800 crew members tested COVID-19 positive.

## Supplementary Tables

**Supplementary Table 1.** Meetings, mostly in an office setting. Infected 9, Susceptible 11, New infections 8

|  |  |  |  |  |  |  |  |  |  |  |
| --- | --- | --- | --- | --- | --- | --- | --- | --- | --- | --- |
| No. | Situation | Place | Occasion | Index patient symptomatic? | Pre- | Susceptible | New Infections | Duration | Distance | Mixing |
| 1. | 1A | Germany | 12 m2 room. Sat on same side of table | N | 1 | 3 | 1 | 1 hour | < 1m* | High |
| 2. | 1C | Germany | Worked together on same computer | N | 1 | 1 | 1 | brief | < 1m | High. |
| 3. | 1D | Germany | Private meeting | N | 1 | 2 | 1 | > 90 min | < 1 m | High |
| 4. | 1E | Germany | Sat back-to-back at canteen and exchanged shaker | N | 1 | 1 | 1 | brief | < 1 m | High, but brief |
| 5. | 1G | Germany | Private meeting | N | 1 | 1 | 1 | 1.5 hr | 1.5 m | High |
| 6. | 1K | Germany | Private meeting | N | 1 | 2 | 2 | 1 hr | 1-1.5 m | High |
| 7. | 18C | USA | Meeting at a church | N | 3 | 1 | 1 | 1.5 hr | 1-1.5 m | High |

**Supplementary Table 2.** Staying together for a few days. Infected 4, Susceptible 15, New infections 8

|  |  |  |  |  |  |  |  |  |  |  |
| --- | --- | --- | --- | --- | --- | --- | --- | --- | --- | --- |
| No. | Situation | Place | Occasion | Index patient symptomatic? | Pre- | Susceptible | New Infections | Duration | Distance | Mixing |
| 1. | 17A | South Korea | Vacation | N | 1 | 3 | 1 | 1 day | - | High |
| 2. | 10 | France | Vacation | N | 1 | 11 | 6 | 4 days | - | High |
| 3 | 11 | Vietnam | Family visit | Y (father was symptomatic) | 2 | 1 | 1 | 3 days | - | High |

**Supplementary Table 3.**
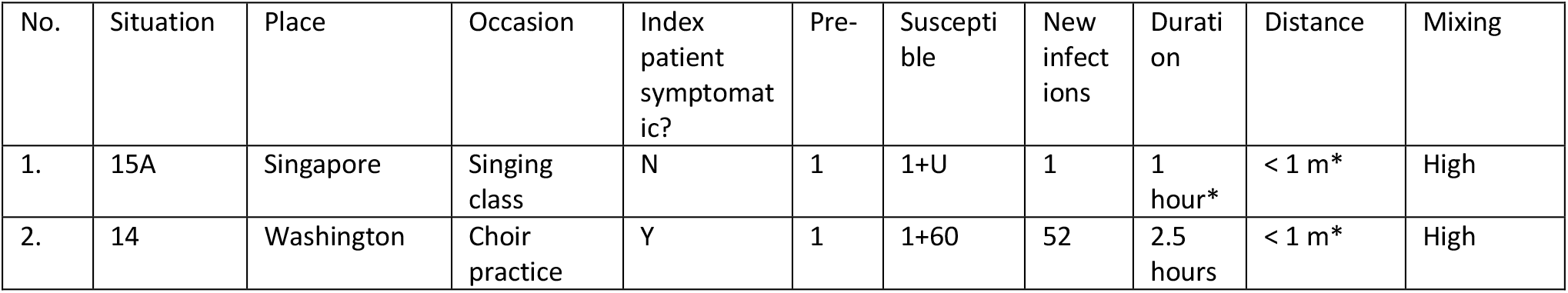
Singing in a closed space. Infected 2, Susceptible 61, New infections, 53

**Supplementary Table 4.**
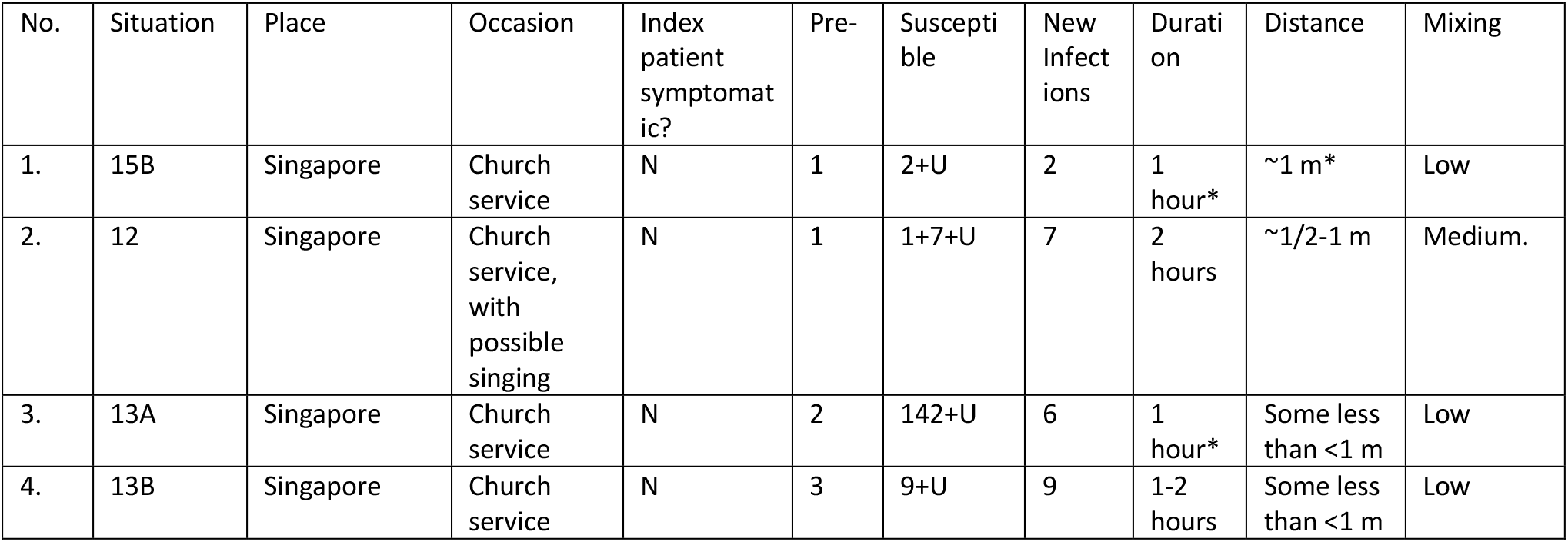
Gathering in a public (closed) space, with possibly no directed talking Infected 7, Susceptible 160, New infections 21

| No. | Situation | Place | Occasion | Index patient symptomatic? | Pre- | Susceptible | New Infections | Duration | Distance | Mixing |
| --- | --- | --- | --- | --- | --- | --- | --- | --- | --- | --- |
| 1. | 15B | Singapore | Church service | N | 1 | 2+U | 2 | 1 hour* | ~1 m* | Low |
| 2. | 12 | Singapore | Church service, with possible singing | N | 1 | 1+7+U | 7 | 2 hours | ~1/2-1 m | Medium. |
| 3. | 13A | Singapore | Church service | N | 2 | 142+U | 6 | 1 hour* | Some less than <1 m | Low |
| 4. | 13B | Singapore | Church service | N | 3 | 9+U | 9 | 1-2 hours | Some less than <1 m | Low |

**Supplementary Table 5.** Shopping and directly interacting with sales agents for an hour. Infected 4, Susceptible 17, New infections 5

| No. | Situation | Place | Occasion | Index patient symptomatic? | Pre- | Susceptible | New Infections | Duration | Distance | Mixing |
| --- | --- | --- | --- | --- | --- | --- | --- | --- | --- | --- |
| 1. | 16B | Singapore | Jewellery shop | Y | 2 | 1+x | 1 | 1 hour | - | High |
| 2. | 16C | Singapore | Health products shop | Y | 2 | 4 if one counts only those who interacted, 16 altogether | 4 | ½ hour | - | High, including touch to apply products |

**Supplementary Table 6.** Family Dinner with strong mixing at and around dinner: Infected 8, Susceptible 30, New infections: 20

| No. | Situation | Country | Occasion | Index patient symptomatic? | Pre- | Susceptible | New infections | Duration | Distance | Mixing |
| --- | --- | --- | --- | --- | --- | --- | --- | --- | --- | --- |
| 1. | 4B | Malaysia | Family dinner | N | 1 | 1+x | 1 | Few hours | - | High |
| 2. | 5 | Singapore | Chinese New Year Dinner | N | 2 | 7+x | 7 | Few hours | - | High |
| 3. | 8 | China | Family dinner | N | 1 | 3 | 3 | Few hours | - | High |
| 4. | 9A | China | Family dinner | N | 1 | 5 | 3 | Few hours | - | High |
| 5. | 9B | China | Family dinner | N | 2 | 11 | 3 | Few hours | - | High |
| 6. | 18A | USA | Dinner at Funeral | Y | 1 | 3 | 3 | 1 day | Close contact | High |

**Supplementary Table 7.** Sit-out Dinner with lesser mixing of attendees: Infected 5, Susceptible 318, New infections: 50

| No. | Situation | Country | Occasion | Index patient symptomatic? | Pre- | Susceptible | New infections | Duration | Distance | Mixing |
| --- | --- | --- | --- | --- | --- | --- | --- | --- | --- | --- |
| 1. | 3B | Singapore | Conference Dining tables | N | 1+1 On two tables | 8+8 On two tables | 2+3 | 3 hours | 1 m | Low since there were events going on |
| 2. | 6 | Singapore | Chinese New Year Dinner | Y | 1 | 200 | 36 | 5 hours | 1 m | Med, there were a few taking pictures |
| 3. | 7A | China | Family dinner at a restaurant | N | 1 | 91 | 9 | 1 hour | 1-2 m | Low in restaurant |
| 4. | 7B | China | Family dinner at a restaurant | N | 1 | 9 or 20 | 9 Same as above | 1 hour | 1-2 m | High along the air conditioning |
| 5. | 1E | Germany | Eating separately, But exchanging salt | N | 1 | 1 | 1 | <1 hour | (< 1m) Within hands reach | High. |

**Supplementary Table 8.**
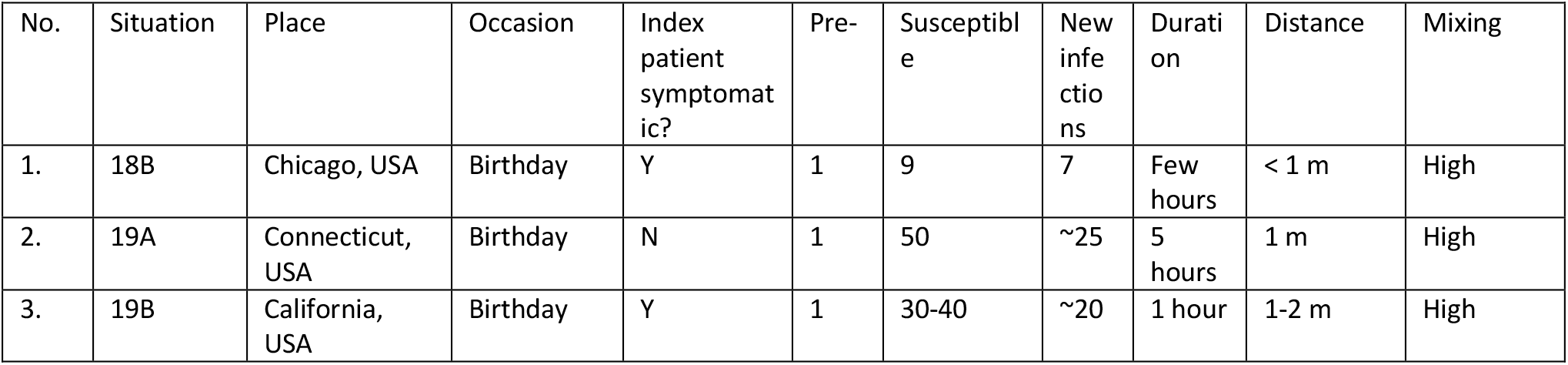
Birthday party. Infected 3, Susceptible 99, New infections: 52

| No. | Situation | Place | Occasion | Index patient symptomatic? | Pre- | Susceptible | New infections | Duration | Distance | Mixing |
| --- | --- | --- | --- | --- | --- | --- | --- | --- | --- | --- |
| 1. | 18B | Chicago, USA | Birthday | Y | 1 | 9 | 7 | Few hours | < 1 m | High |
| 2. | 19A | Connecticut, USA | Birthday | N | 1 | 50 | ~25 | 5 hours | 1 m | High |
| 3. | 19B | California, USA | Birthday | Y | 1 | 30-40 | ~20 | 1 hour | 1-2 m | High |

**Supplementary Table 9.**
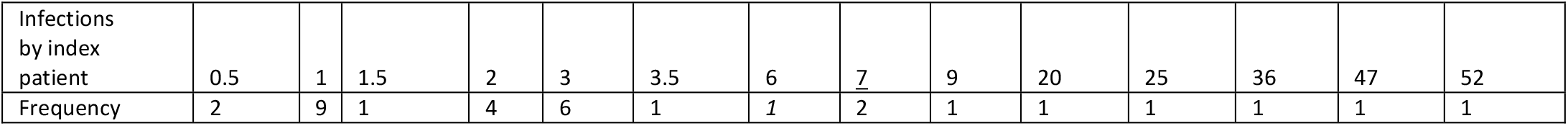
Frequency table of number of infections. Where more than *m* people caused *n* infections, the number of infections per person was considered to be *n/m*.

